# Decoding the endometrial niche of Asherman’s Syndrome at single-cell resolution

**DOI:** 10.1101/2022.10.21.22281346

**Authors:** Xavier Santamaria, Beatriz Roson, Raul Perez, Nandakumar Venkatesan, Javier Gonzalez-Fernandez, Estefania Fernández, Inmaculada Moreno, Hugo Vankelecom, Felipe Vilella, Carlos Simon

## Abstract

Asherman’s Syndrome (AS) is characterized by intrauterine adhesions, which cause infertility, menstrual abnormalities, and recurrent pregnancy loss. While AS occurs as a consequence of traumatic or infectious disruption of the endometrial cell niche, its pathophysiology remains largely unknown and treatment strategies have been restricted to recurrent hysteroscopic removal of intrauterine adhesions with limited success.

We decoded the disrupted endometrial cell niche associated with AS at single-cell (sc) resolution by analyzing transcriptomic data from over 230,000 cells. We sought to prove the functional relevance of our findings by incorporating scRNA-seq analysis into a phase I/II clinical trial of CD133+ bone marrow-derived stem cells in AS patients (EudraCT Number: 2016-003975-23) and through in vitro analysis of AS patient-derived endometrial organoids.

Our integrated analyses supported the construction of an atlas describing the dysfunctional endometrial niche of AS patients, characterized by significant differences in cell population ratios, differential gene expression, and aberrant cell-to-cell communication. Our AS atlas also highlights the existence of two unique cell types – a stressed epithelial population (AS epithelium) expressing the secretory leukocyte protease inhibitor (SLPI) and a population of smooth muscle cells expressing ACTG2 (SMC). These alterations act together to maintain a dysfunctional pro-fibrotic, pro-inflammatory, and anti-angiogenic environment; however, we describe the partial reversion of the cellular, transcriptomic, and aberrant cell-to-cell communication differences *in vivo* and *in vitro* (using endometrial organoids) by patient-specific cell therapy.

This first description of a comprehensive functional endometrial cell atlas of AS provides a holistic view of the disrupted AS-associated endometrial niche, thereby providing insight into pathophysiology and aiding the development of advanced therapeutics.

## Introduction

Asherman’s Syndrome (AS) is an acquired pathological condition defined by the presence of intrauterine adhesions (IUAs) that cause the uterine walls to adhere to each other, resulting in menstrual abnormalities, pelvic pain, infertility, recurrent miscarriage, and abnormal placentation^1^. AS affects 4 in every 10,000 women^2^ and is considered a rare disease (See Orphanet database; ORPHA:137686). Trauma or infection induced by procedures such as curettage or cesarean section, mainly in a gravid uterus, can disrupt the endometrial basalis and prompt AS onset^3–6^.

Since the first report of AS as traumatic amenorrhea more than a century ago^7,8^, treatment strategies have focused on the recurrent hysteroscopic removal of IUAs with preventative measures meeting with limited clinical results, especially in the moderate and severe stages refractory to surgical treatment^9,10^. IUAs do not cause AS but instead occur due to the iatrogenic disruption of the endometrial niche^11^. Although bulk tissue-based transcriptomics studies^12–14^ have revealed basic information, convincing evidence regarding the molecular mechanisms involved in AS-related endometrial dysfunction remains elusive. Advancements in single-cell RNA sequencing (scRNA-seq) of the human endometrium^15^ and endometrial organoid culture systems^16^ have enabled a deeper understanding of cellular cartography and cell-to-cell communication (CCC) in endometrial pathologies such as endometriosis^17^ and atrophic endometrium^18^.

Deciphering the cellular complexity and heterogeneity of the endometrial cell niche will provide a means to better understand the etiological causes of IUAs formation and endometrial dysfunction in AS. For this purpose, we performed systematic differential cellular, transcriptomic, and immunological comparisons at single-cell resolution and sought to explore differential CCC within the normal and AS endometrium. Furthermore, we also evaluated the impact of patient-specific autologous cell therapy (CD133+ bone marrow-derived cells; BMDSCs) in vivo and in matched AS patient-derived endometrial organoids in vitro at the single-cell level to prove the functional relevance of our findings.

Our data highlight significant differences between the endometrial atlases of healthy and AS patients, consisting of the presence of two unusual cell types, significant differences in cell population ratios, differential gene expression profiles, and aberrant CCC. Our results support the existence of a dysfunctional pro-fibrotic, pro-inflammatory, and anti-angiogenic endometrial environment associated with AS that can be partially reverted by patient-specific cell therapy. Overall, these findings provide a platform for a deeper understanding of AS, which may support the development of improved preventative and therapeutic approaches.

## Results

### Single-cell analysis reveals a differential transcriptomic profile and cellular cartography in the endometrium of human AS patients

scRNA-seq was initially applied to characterize the differential endometrial cartography of AS through a comparison of a total of 110,022 cell transcriptomes (**Fig. 1**). Secretory-phase endometrial biopsies were collected by hysteroscopy from 9 individuals with moderate and severe AS according to the American Fertility Society classification of uterine adhesions^19^ (**Methods** and **Supplementary Data**). A total of 69,202 single-cell transcriptomes from 10 fertile women as controls of normal secretory endometrium were analyzed (available at GSE111976 dataset^15^). We generated a comparative cellular landscape of the AS dysfunctional endometrium using an integration pipeline (**Methods**). Then, clustered cells were visualized in uniform manifold approximation and projection (UMAP) coordinates after dimensional reduction. Subsequently, cell types were identified by exploration of cluster marker genes (**Supplementary Tables 1** and **2**) that were compared to annotated major cell types in reported endometrial atlases^15,20^. Fine-tuning of subpopulations was performed by manual curation of markers.

**Figure 1.**
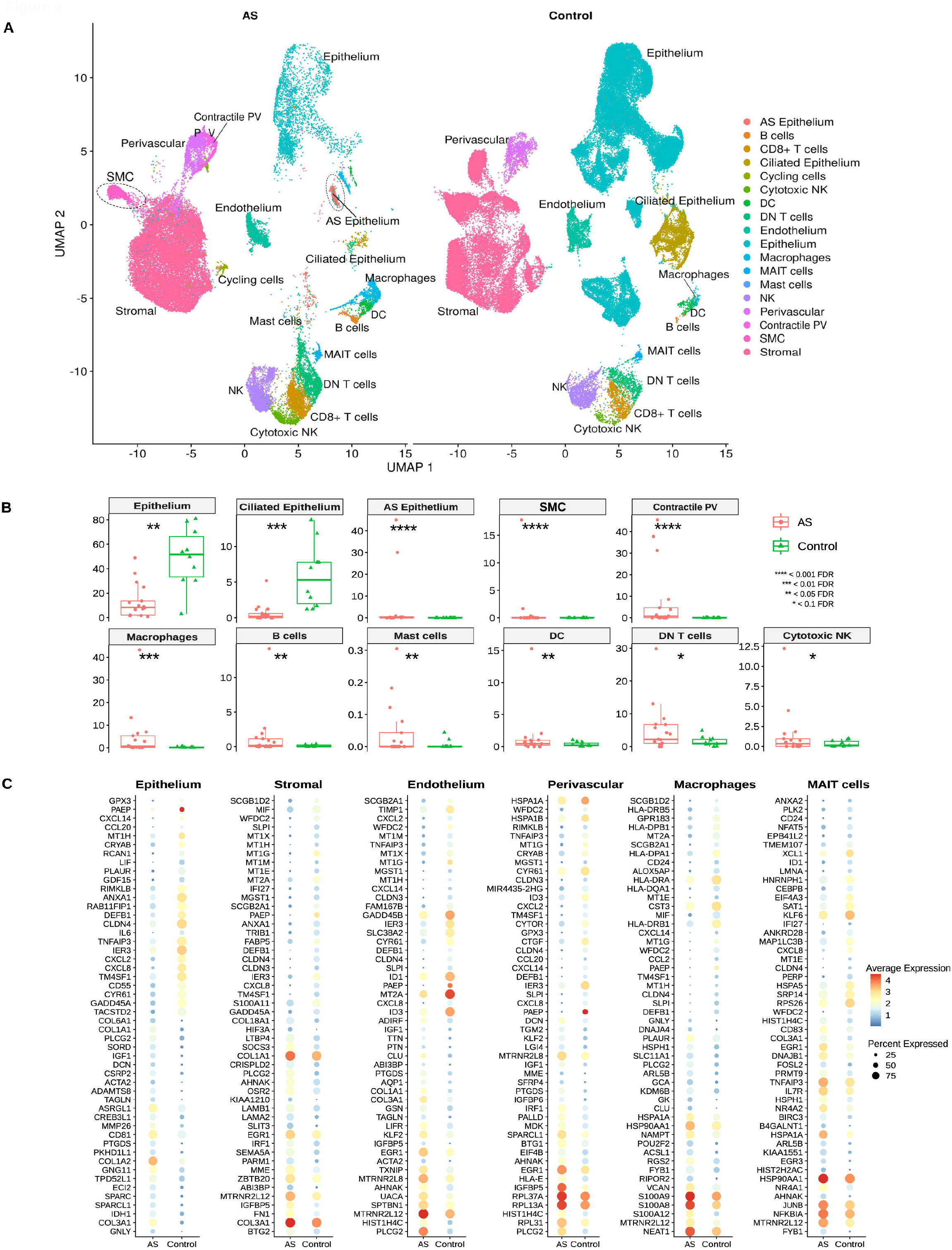
scRNA-seq cartography of the human endometrium in Asherman’s syndrome patients and healthy controls. **A)** UMAP (Uniform manifold approximation and projection) integration of 109,538 cells from nine Asherman’s Syndrome (AS) patients (N=40,336 cells) and ten healthy individuals (Control) from the GSE111976 dataset (N=69,202 cells). Unusual AS-related cell types are highlighted within dotted circles (SMC cells and AS Epithelium). **B)** Cell ratios comparing the endometrium of healthy (Control) and AS patients. **C)** Differentially expressed genes when comparing AS and Control endometria by specific cell types. Abbreviations: DC: dendritic cells; DN: double negative; AS: Asherman; MAIT: mucosal-associated invariant T; NK: natural killer; PV: perivascular; SMC: smooth muscle cells; FDR: false discovery rate.

We identified eighteen cell types in our integrated AS endometrial atlas (**Fig. 1A)**. Stromal fibroblasts expressed canonical markers (DCN, LUM, and IGF1), epithelial cells expressed members of the metallothionein gene family (MT1G, MT1H, MT1X, and MT1M) and WFDC2, while ciliated epithelial cells expressed genes related to the formation and maintenance of cilia (C20orf85, C1orf194, and C9orf24). Two identified perivascular (PV) cell clusters expressed the regulator of G protein signaling gene (RGS5) and genes related to microfilaments (ACTA2, TAGLN, and MYL9). One PV cell cluster displayed a more robust expression of ADIRF and genes involved in contractile functions (ACTA2 and MHY11); therefore, we labeled this cell subtype contractile PV. Endothelial cells expressed A2M and secreted proteins such as VWF and ENG (a glycoprotein involved in the structure and integrity of the vasculature). The myeloid lineage cell cluster expressed classic macrophage markers (e.g., LYZ) and genes encoding S100 calcium-binding proteins (S100A8 and S100A9), while dendritic cells expressed regulatory antigen processing CD74 and HLA class II genes. We also identified the following lymphoid lineage cell clusters: natural killer (NK) cells by NKG7 gene expression; cytotoxic NK cells by serine protease (GZMH and GZMB) and cytolytic related gene (PRF1) expression; CD8+ T cells by IFNG, IL32, and CCL5 gene expression; and a population of double-negative (DN) T cells by IL7R, LTB, CD52, and CD3D expression. A specific population of innate-like T cells - defined by their semi-invariant αβ T cell receptor (TCR) denominated as mucosal-associated invariant T (MAIT) cells^21^ – was also identified through the specific expression of IL23R, IL4I1, LTB, IL7R, and KLRB1. These genes provide an innate capacity to respond to a specific set of ligands without needing to expand and develop memory T cells (**Fig. 1A** and **Supplementary Fig. 1A-C**).

We found significant differences in cell population ratios between AS and control endometrial samples (**Fig. 1A-B**). Importantly, we discovered a significant reduction in the epithelium of the AS endometrium compared to healthy control (8.3% vs. 51.65%), which primarily impacted ciliated epithelium (0.19% vs. 5.3%). Myeloid and lymphoid cell lineages displayed more abundance in the AS endometrium than in the healthy control, which included a highly significant increase in macrophages (0.6% vs. 0.08%), B cells (0.13% vs. 0.015%), dendritic cells (0.45% vs. 0.18%), DN T cells (2.22% vs. 0.94%), and cytotoxic NK cells (0.32% vs. 0.11%). We also observed an increased abundance of contractile PV cells in the AS endometrium compared to the healthy control (0.74% vs. 0.01%) (**Fig. 1B**).

Beyond the expected endometrial cell populations, we identified two unique cell types in AS (**Fig. 1A-B**). These comprised a stressed epithelial subpopulation (AS Epithelium) that displayed high levels of SLPI gene expression, which encodes immunoprotective protein secretory leukocyte protease inhibitor, and co-expressed cell stress-related genes (i.e., HSPA1A and SOCS3) and smooth muscle cells expressing ACTG2 (SMC).

Importantly, we found differential gene expression in several cell compartments in AS compared to the healthy control endometrium (**Fig. 1C** and **Supplementary Fig. 2**). In the epithelium, we fail to find expression of critical genes involved in the opening of the window of implantation (WOI), including PAEP, GPX3, and CXCL14, and members of the metallothionein (MT) family. Interestingly, we observed the upregulated expression of pro-fibrotic (collagen family genes and FN1) and anti-angiogenic genes (ADAMTS8 and DCN)^22^ in the AS endometrium. Similarly, AS stromal cells displayed an upregulation of pro-fibrotic genes (collagens, FN1, and the laminins LAMB1 and LAMA2), the extracellular matrix (ECM) anchor ABI3BP^23^, and an anti-angiogenic factor (IGFBP5)^24^ AS stromal cells displayed a lower expression of secretoglobin genes (SCGB1D2 and SCGB2A1) involved in the secretory transformation of the menstrual cycle. Both the perivascular and endothelial cell types (**Fig. 1C**) displayed the upregulated expression of genes associated with the stress response to endothelial injury, including CLU^25^, KLF2^26^, and EGR1^27,28^ and the anti-angiogenic gene factors IGFBP5 and IGFBP6^24,29^. We uncovered a potential aberration in endometrial permeability in AS, as shown by the significant downregulation of tight junction-related genes such as CLDN4 and CLDN3 in the endothelium and the non-contractile perivascular population. Our data also provided evidence for a global pro-inflammatory, activated immune cell state in the AS endometrium. Macrophages overexpressed S100 family genes (S100A8/9/12)^30^ and the long non-coding (lnc) RNA NEAT1, which has been associated with inflammation and immune activation^31^. MAIT cells presented a differential gene expression signature compared to controls through the more robust expression of early growth response transcription factors EGR1 and EGR3, two hub gene expression re-programmers JUNB and FOSL2, and the TCR signal–associated nuclear hormone receptor NR4A1 (**Fig. 1C**).

Together, these data show that the AS endometrium is characterized by dysfunctional epithelium, fibrotic stroma, a pro-inflammatory polarized microenvironment with disrupted angiogenesis and a compromised permeability in blood vessels.

### Aberrant cell-to-cell communication in AS underlies the dysfunctional profibrotic, pro-inflammatory, and anti-angiogenic environment

Cell-to-cell communication (CCC) plays a critical role in maintaining tissue homeostasis. To dissect the complex interactions between endometrial cell types and their possible interruption in AS patients, we inferred all potential intercellular communications by analyzing the expression of ligand-receptor pairs using CellChat^32^. We calculated communication information flow, defined as the total interaction probability, among all pairs of cell populations present in AS and controls and analyzed differential signaling for each pathway (**Fig. 2** and **Supplementary Fig. 3A**).

**Figure 2.**
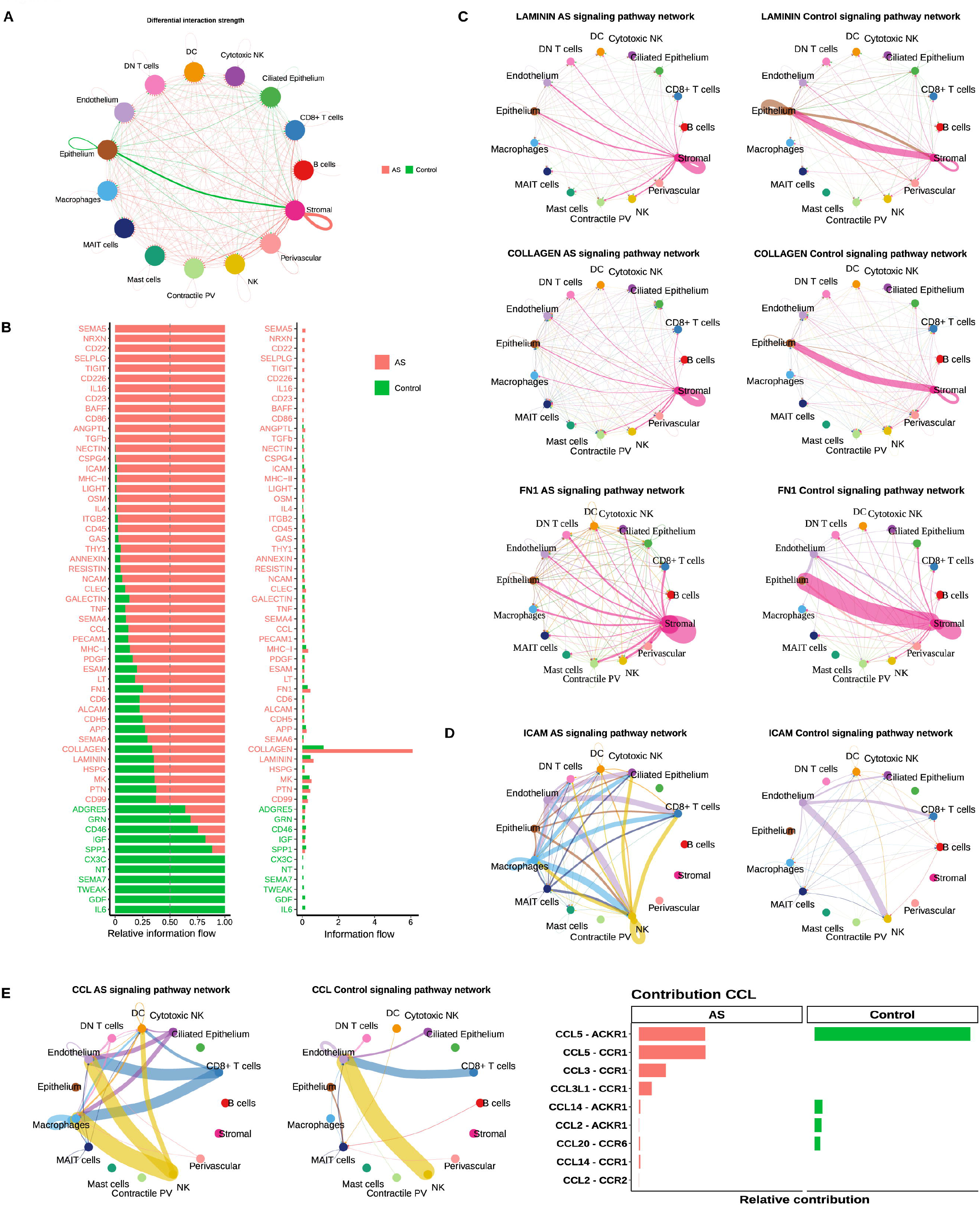
Differential cell-to-cell communications (CCC) between the human endometrium in Asherman’s syndrome patients and healthy controls. **A)** Chord plot displaying differentially active CCCs between AS and control groups. Arrows between cell types show the direction of interactions. And the relative thickness of each red (AS) and green (Control) arrow lines represents the expression-based strength of the interaction between cell types. **B)** Relative and absolute information flows of each signaling pathway differentially detected between AS and control groups. **C)** CCC chord plots of LAMININ, COLLAGEN, ICAM and CCL signaling pathways in the AS and Control endometria. Arrows follow the color code of cell types commonly detected in AS and Control cartographies. **D)** Ligands and receptors contributing to CCCs of the C-C motif chemokine ligand (CCL) signaling pathway.

Our first view of CCC networks demonstrated a significant loss of normal interactions between epithelial and stromal cells, which shifted to autocrine stimulation of the stromal population (**Fig. 2A**). The upregulation of ligands and receptors from collagen, FN1, and laminin ECM-related pathways in the AS endometrium (**Fig. 2B** and **Supplementary Fig. 3)** supported this increase in autocrine stromal signaling (**Fig. 2C** and **Supplementary Fig. 4-6**), which is consistent with the pro-fibrotic nature of AS. We also identified increased number and intensity of signaling interactions between immune cells in AS through ICAM and CCL (**Fig. 2D-E**, and **Supplementary Fig. 7**), suggesting pro-inflammatory and cell recruitment signaling pathways, respectively. We observed an enrichment of signaling pathways related to major immune regulatory processes, such as ICAM, MHC-II and MHC-I, LIGHT, IL4, RESISTIN, NCAM, and CCL (**Fig. 2B** and **Supplementary Fig. 3B)**. The AS endometrium also displayed increased communication between endothelium and immune cells (macrophages, NKs cells, and T cell populations) via ICAM (**Fig. 2D**) and integrin pathways (**Supplementary Fig. 3B**)^33^.

Therefore, our results uncover a marked dysfunction in CCC and cell-to-matrix interactions along the stromal-epithelial axis leading to a pro-fibrotic stromal subpopulation in AS. In parallel, blood vessels facilitate leukocyte/myeloid extravasation to the tissue attracted by the chemokine-rich pro-inflammatory environment.

### Partial reversion of abnormal cellular and transcriptomic profiles in AS endometrium following patient-specific autologous cell therapy

The European Medicines Agency (EMA) (2017)^2^ and the Food and Drug Administration (FDA) (2018) (https://www.accessdata.fda.gov/scripts/opdlisting/oopd/detailedIndex.cfm?cfgridkey=613117) approved autologous cell therapy using CD133+ BMDSCs^34^ as the first orphan drug designated product for AS treatment. This Advanced Therapy Medicinal Product (ATMP) is currently in phase I/II trials (EudraCT Number: 2016-003975-23; Registration date: April 21, 2021). Thus, we incorporated scRNA-seq analysis into this clinical trial in AS patients to explore the functional relevance of the AS endometrial cell atlas.

We analyzed endometrial biopsies from each patient before and after administration of CD133+ BMDSCs to determine responses to cell therapy (**Supplementary Fig. 8**). A total of 123,250 AS patient-derived cells (pre-treatment n=40,336 cells; post-treatment n=82,914 cells) were assessed. We integrated matched pre-treatment and post-treatment scRNA-seq samples into the same space of reduced dimensions (**Fig. 3A**) and then compared cell types by changes in cell ratios (**Fig. 3B** and **Supplementary Fig. 9**) and differential gene expression (**Fig. 3C-D** and **Supplementary Fig. 10**). The administration of autologous cell therapy to AS patients prompted an increase in the epithelial compartment compared to pre-treatment (from 8.3% to 10%), with a statistically significant reduction in the disease-specific AS epithelium population (0.035% vs. 0.017%, respectively). Moreover, we found a significant reduction in the macrophage population (0.61% vs. 0.21%) (**Fig. 3B**).

**Figure 3.**
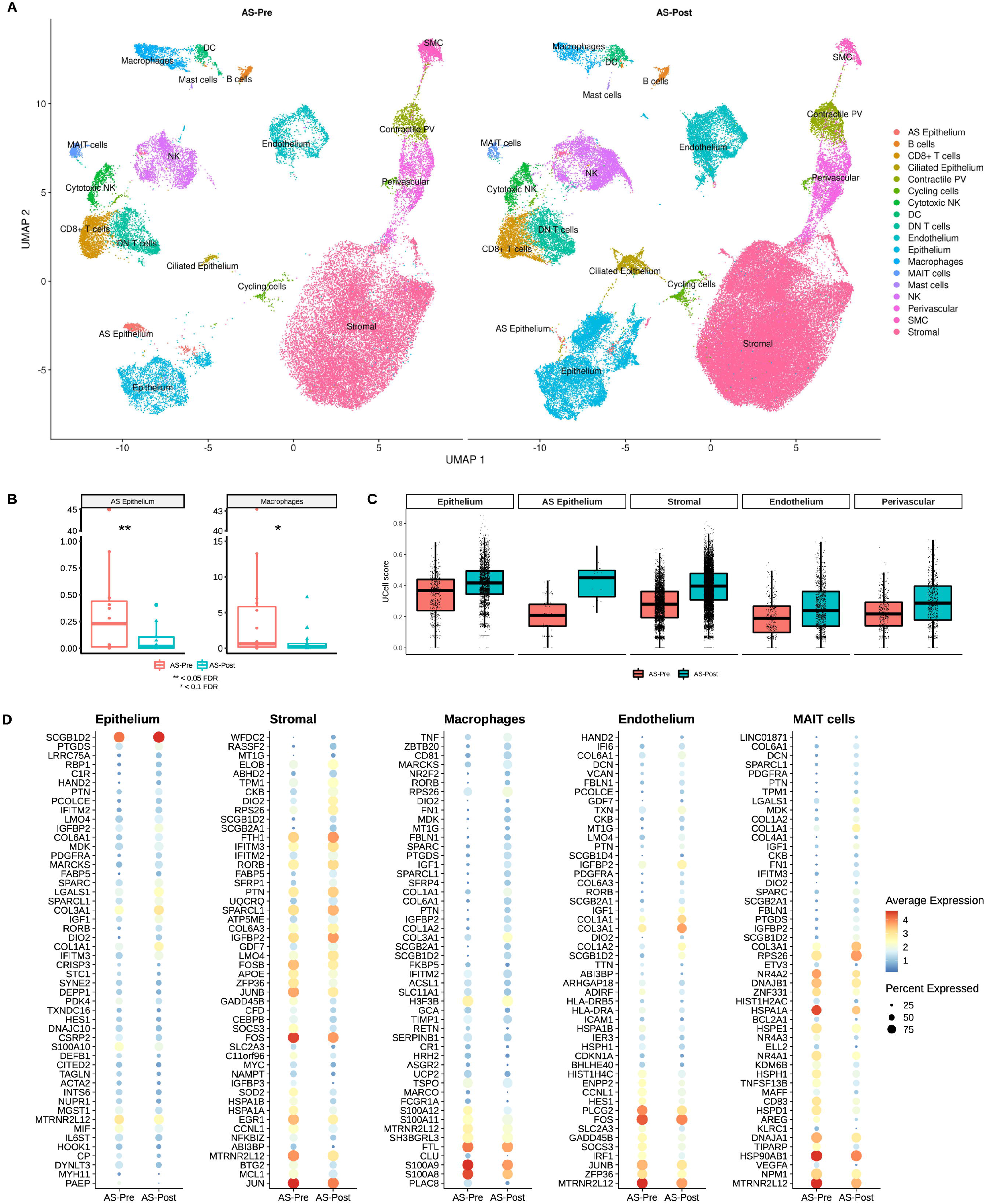
Transcriptional analysis in AS patients pre-treatment and post-treatment with CD133+ BMDSC therapy. **A)** UMAP of scRNA-seq results from nine AS patients at pre-treatment (AS-Pre: N=40,336 cells) and post-treatment stage (AS-Post: N=82,914 cells). **B)** Boxplot of cell type ratios comparing AS patients at pre-treatment (AS-Pre) and post-treatment (AS-Post) stages. **C)** Boxplot displaying the results of the ssGSEA enrichment over a gene set of the secretory phase of the menstrual cycle. **D)** Dotplot of the differential expressed genes comparing AS patients at pre-treatment (AS-Pre) and post-treatment (AS-Post) stages in the epithelium, stroma, endothelium and macrophages.

Differential gene expression analysis prior and following cell therapy in AS patients revealed significant transcriptomic changes in endometrial cell populations (**Fig. 3D, Supplementary Fig. 10**, and **Supplementary Table 5**). In the epithelium, genes related to epithelial development and cell proliferation were upregulated. These included IGF1^35^ and PDGFRA, which plays a pivotal role in ECM remodeling and angiogenesis^36^ after BDMSC administration. We also observed an increase in the expression of secretoglobin genes after BDMSC administration, which play essential roles in the secretory activity of glands^15,37^. Finally, we observed the downregulated expression of genes related to the immune response and chronic inflammation after BDMSC administration, which included MIF, IL6ST, DEFB1, and S100A10. S100A10 promotes macrophage recruitment^38^, while DEFB1 is a host defense peptide with antimicrobial and pro-inflammatory properties^39^.

In the stromal compartment, cell therapy induced the recovery of gene expression profiles related to the normal functioning of the menstrual cycle in the secretory phase (e.g., SCGB1D2, SCGB2A1, and MT1G) (**Fig. 3D**). Stromal cells in the post-treatment group also displayed a downregulation of the cellular stress genes HSPA1A and HSPA1B, oxidative stress response (SOD2), and fibrosis (BTG2), the overexpression of pro-angiogenic and vascular development factors in the post-treatment group, including IGFBP2^29^ and the downregulation of IGFBP3, which is related to anti-angiogenic processes under pro-inflammatory environments^40^.

In the endothelial fraction (**Fig. 3D**), BDMSC administration induced a significant transcriptomic change from a stressed/pro-inflammatory profile to a more homeostatic profile. Changes included the downregulation of genes related to antigen presentation (HLA-DRB5 and HLA-DRA) and leukocyte/myeloid trans-endothelial migration (ICAM) and heat shock proteins (HSPA1B and HSPH1). We also observed the overexpression of genes related to angiogenesis (IGFBP2)^29^ and ECM and cell adhesion (COL1A1, COL3A1, COL6A3, COL6A1, and VCAN). Collagen types I and III in the vessel surroundings have a pivotal role in capillary morphogenesis to guide new vessel sprouts^41^.

After cell therapy, endometrial macrophages downregulated genes related to pro-inflammatory and adaptive immune responses, such as the S100A family genes (**Fig. 3D)**. Although we did not observe a significant change in the population ratios of MAIT cells (0.16% vs. 0.09), the modulated expression of heat-shock protein genes in MAIT cells suggested the existence of a lower stress profile in post-treatment AS patients (**Fig. 3D**).

Regarding the restoration of endometrial function, Gene Set Enrichment Analysis (GSEA) analysis yielded a significant enrichment in genes typically expressed during the secretory phase of the menstrual cycle in post-treatment AS patients in the epithelium, stroma, endothelium, and perivascular cell fractions, compared to the pre-treatment stage (**Fig. 3C)**.

Combined, these findings show that cell therapy reverts the AS epithelium population and macrophages to a healthy-like state, induces the recovery of functionality in major cell types characterized by a reduction in fibrosis, oxidative stress, and inflammation, and triggers an increase in angiogenesis.

### Cell-to-cell communication in the AS endometrium in response to cell therapy

CCC network analysis in AS patients after cell therapy reported the enrichment of major signaling pathways, including GDF, TWEAK, CX3C, and IGF pathways **(Fig. 4A**), which induced the appearance of a healthy-like profile **(Fig. 2B)**. Meanwhile, we observed a reduction in pro-inflammatory and immune regulation for the RESISTIN, IL4, CCL, CXCL, ANGPTL, MHC-I, MHC-II, and LIGHT signaling pathways in AS patients after cell therapy **(Fig. 4A**). We also observed significant changes in signaling pathways related to angiogenesis and fibrosis. In the fibroblast growth factor (FGF) signaling pathway **(Fig. 4B)**, ligands had different contributions to the interactions. FGF7 and FGF10 displayed prevalence in the post-treatment group (**Supplementary Fig. 11)**, with FGF7 activating epithelial proliferation and survival and is considered a potential therapy for lung fibrosis^42^ and FGF10 promoting endothelial growth and angiogenesis in corneal wounds^43^. FGF2 was the only ligand detected at the pre-treatment stage. In the post-treatment group, a decrease in the ICAM pathway occurred alongside lower communication flow between endothelial cells and macrophages, NK cells, DN T cells, CD8+ T cells, and DC cells (**Supplementary Fig. 12A)**. We also observed activation of the WNT signaling pathway, supporting increased communication between the stroma and epithelium (**Fig. 4C**). Interestingly, NRG signaling activates after cell therapy (**Fig. 4D**), leading to cell proliferation and differentiation after being released by stromal cells and binding ERBB receptor in epithelial cells^44^. In the angiogenic ANGPTL signaling pathway, the ANGPTL2 ligand prevails after cell therapy (**Fig. 4E**). The co-existence of ANGPTL1 and ANGPTL2 modulates angiogenesis and vascular regeneration processes^45^. Finally, chronic inflammation-associated LIGHT signaling decreased strongly (**Supplementary Fig. 12B**), while IL4 and RESISTIN signaling pathways became inactivated post-treatment (**Supplementary Fig. 12C-D**).

**Figure 4.**
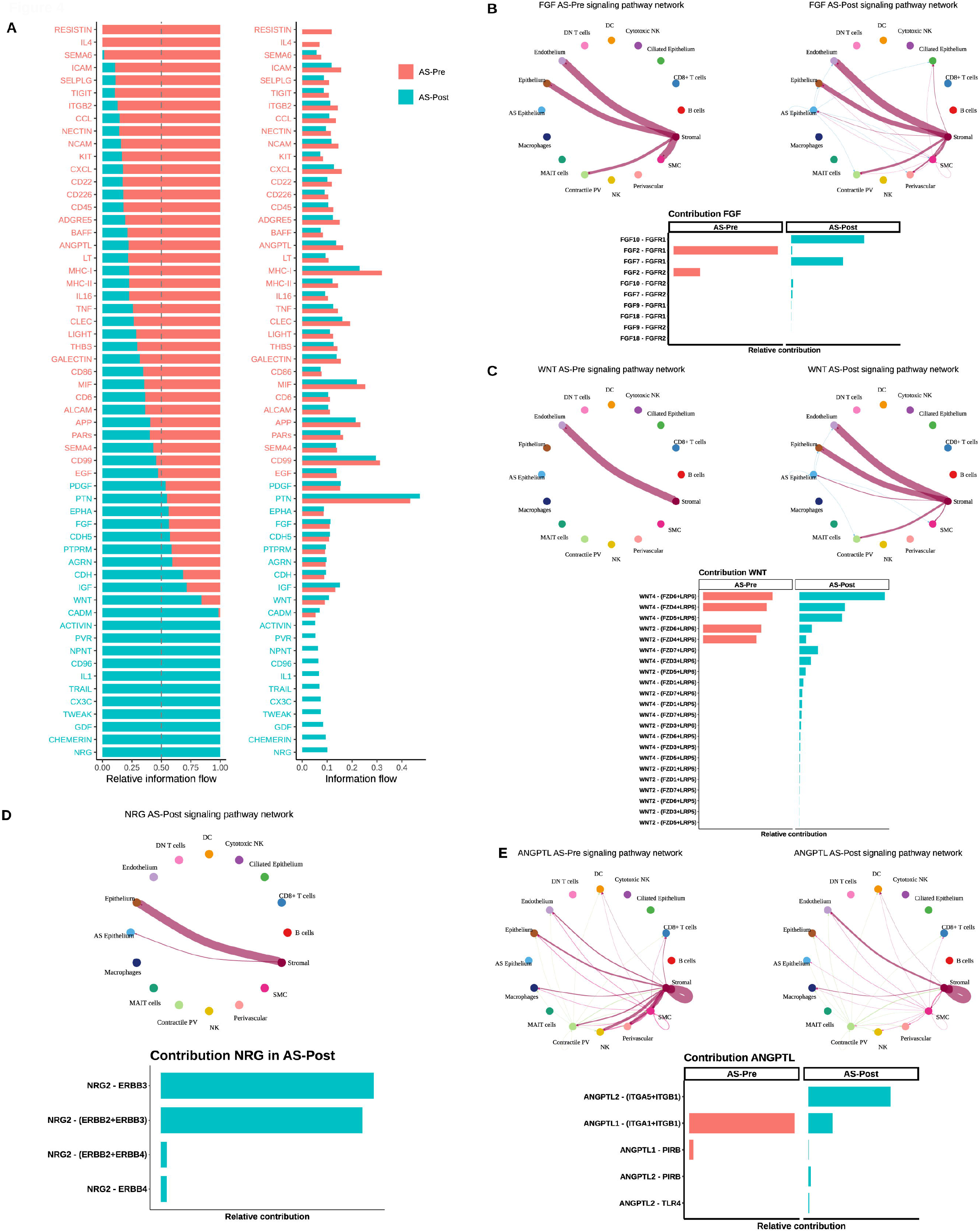
Changes in cell-to-cell communication (CCC) in the AS endometrium pre-treatment and post-treatment with CD133+ BMDSC therapy. **A)** Relative and absolute information flows of each signaling pathway differentially detected in AS patients between pre-treatment (AS-Pre) and post-treatment (AS-Post) stages. Blue dots represent immune system signaling pathways. Red dots represent pathways related to vascular and angiogenesis processes. Green dots represent pathways involved in epithelium development. **(B-E)** For each signaling pathway, CCC chord plots (upper panels), and pathway interactor molecules (lower panel) in AS-Pre and AS-Post stages. **B)** FGF signaling pathway. **C)** WNT signaling pathway. **D)** NRG signaling pathway. **E)** ANGPTL signaling pathway.

Therefore, cell therapy in AS patients activates angiogenic, proliferative, and cell differentiation signaling pathways to the detriment of pro-inflammatory pathways; specifically, GDF, TWEAK, CX3C, and IGF pathways returned to a healthy-like state.

### AS patient-derived organoids recapitulate AS and the effect of cell therapy

To further evaluate our *in vivo* results regarding the dysfunctional pro-fibrotic, pro-inflammatory, and anti-angiogenic endometrial environment of AS in the epithelial compartment and their partial reversion by patient-specific cell therapy, we profiled endometrial organoids at the single-cell level to benchmark this model system against our *in vivo* data. We obtained endometrial cell samples from 3 AS patients pre-treatment and post-treatment and 3 healthy controls. We then generated epithelial organoids and passaged them twice to remove contaminating stromal cells (**Supplementary Fig. 13A)**. We observed an increase in the number of organoid-forming potential in cells isolated from post-treatment (P0 and P1) AS patients compared to cells isolated from pre-treatment AS patients. An equilibrium in organoid formation pre- and post-treatment was reached at P2, indicating a positive selection of organoid-forming cells during *in vitro* culture (**Supplementary Fig. 13B-C**).

Single-cell analysis of organoids yielded 38,162 high-quality cells (pretreatment n=16,985, post-treatment n=13,890, and controls n=7,287). We employed single-cell transcriptomic profiles to classify organoid cell identities, inferring them from *in vivo* data by a logistic regression predictive model. We projected recovered cells in individual UMAPs (**Fig. 5A**). Four main subpopulations of previously described epithelial cells were identified based on canonical markers: luminal (PTGS1), glandular (PAEP and GPX3), ciliated (TPPP3 and C11orf88), and SOX9+ cells^15,20^ (**Fig. 5B** and **Supplementary Fig. 13D**).

**Figure 5.**
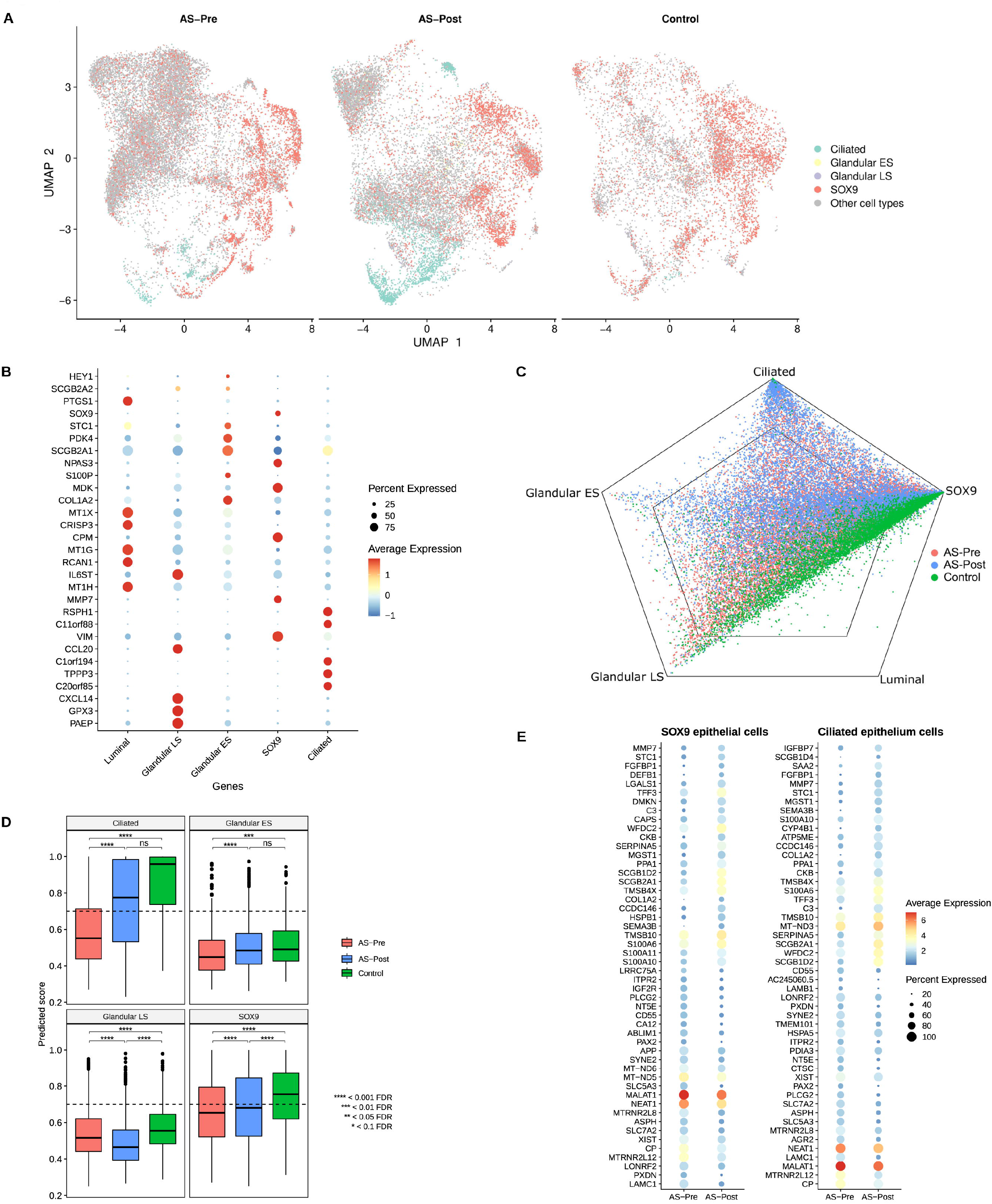
Analysis of endometrial epithelial organoids derived pre-treatment and post-treatment from AS patients with CD133+ BMDSC Therapy and healthy controls. **A)** UMAP of the main cell types used to train the logistic regression machine learning model. Cell types were labeled in the epithelial fraction of a healthy data subset. Organoid dataset and the inferred cell types by logistic regression model. Cells with a predicted score < 0.7 were labeled as “Other cell types”. **B)** Dotplot displaying the expression of marker genes for each cell type. **C)** Polygonal representation of cell type probabilities predicted by a logistic model trained on epithelial sub-cell types in vivo. **D)** Boxplot of each cell type per organoid group. Dunn test was applied to evaluate the differences between groups. p-value adjusted by Bonferroni method. **E)** Dotplot displaying the top 50 genes differential expressed in SOX9 and ciliated cells population in the pre-treatment (AS-Pre) and post-treatment (AS-Post) groups. Abbreviations: Early Secretory; LS: Late Secretory.

We employed the logistic regression model as the similarity score between the *in vivo* and *in vitro* systems to measure the differences between pre-treatment and post-treatment organoids. Regarding cell identity, a polygonal plot shows the highest similarity scores between organoids and their *in vivo* counterparts for epithelial ciliated and SOX9+ cell subtypes (**Fig. 5C**), while lower scores were observed for the glandular cells of early (ES) and late secretory (LS) subtypes (**Fig. 5C**). The molecular identity of epithelial ciliated, SOX9+, and ES glandular cells derived from post-treatment organoids became progressively similar to control organoids and in-vivo healthy counterparts (**Fig. 5D** and **Supplementary Fig. 13D-E)**. Finally, differential gene expression analysis in the SOX9+ and ciliated cell populations revealed down-regulation of LAMC1 (a specific component of the basal ECM) in post-treatment organoids compared to pre-treatment organoids (**Fig. 5E**). In parallel, two humanin micropeptides derived from the mitochondria (MTRNR2L8 and MTRNR2L12) and two lncRNAs (NEAT1 and MALAT1) closely related to multiple inflammatory disorders ^31^ became downregulated in post-treatment organoids.

Together, these results suggest that endometrial organoids represent a robust *in vitro* model for AS syndrome. Further, cell therapy promotes cellular and transcriptomic changes that partially resemble control organoids.

## Discussion

We report a comprehensive functional single-cell AS endometrium atlas, which identifies 18 cell types - two uniquely present in AS – and describes significant differences in cell population ratios, gene expression and CCC that support a dysfunctional pro-fibrotic, pro-inflammatory and anti-angiogenic endometrial environment (**Fig. 1**).

AS epithelial cells and SMC cells represent two disease-associated cell types^46,47^. The AS-epithelium overexpressed genes related to cellular stress responses (HSPA1A) and immune response regulators such as SOCS3, an intrinsic protein that controls STAT3 activation by JAK inhibition^48^. Their overexpression in colon epithelial and lamina propria cells in bowel inflammatory disease and skin epithelial cells suggests importance to inflammatory control^49^ and wound healing^50^.The main sign of AS is the presence of fibrosis in the form of IUAs. In fibrosis-a reactive process that develops in response to excessive epithelial injury and inflammation-, fibroblasts synthesize greater amounts of collagens and proteoglycans raising the stiffness of the tissue, which causes further myofibroblast activation and epithelial damage^51^. ACTG2-expressing SMCs, together with fibroblasts, have been identified in the development of idiopathic pulmonary fibrosis^52–55^. Histologically, IUAs are mostly composed of collagen bundles, although some muscular fibers can be found in the central part of the adhesion bridge in the most severe cases^56^. In our study, we only encountered this unique cell type in moderate and severe AS, but absent following cell therapy, indicating the functional relevance of our findings because it could be reverted.

We found significant differences in cell population ratios in the AS endometrium, including a prominent reduction in the epithelial population, especially the ciliated epithelium that is consistently present in healthy endometrium^15^. Disease-associated alterations occurred alongside the downregulation of WOI-associated genes (e.g., PAEP, GPX3, CXCL14, and MT) and the upregulation of pro-fibrotic and anti-angiogenic genes (e.g., ADAMTS8 and DCN), which may underly the dysfunctional receptive epithelium implicated in the poor reproductive outcomes of AS patients^57^. AS-associated stroma upregulated pro-fibrotic genes (collagens, FN1, LAMB1, and LAMA2), the ECM anchor ABI3BP gene^23^, and genes coding anti-angiogenic factors (such as IGFBP), which may be implicated in the altered invasion phase of abnormal placentation^1^. AS-specific increases in myeloid/lymphoid cell lineages represented by macrophages, CD8+ T cells, double-negative T cells, NK cells, and cytotoxic NKs cells indicate a pro-inflammatory state. Macrophages overexpressed S100 family genes (S100A8/9/12)^30^ and the lncRNA NEAT1, while DCs overexpressed the apoptotic inhibitor MCL1 and the antimicrobial pro-inflammatory peptide GNLY, which also becomes dysregulated in CD8+ T cells, DN T cells, and NK cells. Myeloid populations, T cells, NK cells, and cytotoxic NK cells from AS patients express PLCG2, which encodes a signaling enzyme essential for immune system receptors.

Analyses of ligand-receptor pair expression in AS revealed a CCC shift from epithelial-stromal interactions to stromal autocrine signaling, supporting the pro-fibrotic nature of AS. The endothelium becomes permeable to leukocyte recruitment and transendothelial migration through the ICAM pathway, producing pro-inflammatory cytokines (e.g., CCL5, CCL3, and CC3L1) by CD8+ T cells and NK cells targeting CCR1 receptors in macrophages and DCs supporting leukocyte/myeloid cell recruitment. The CCL5-CCR1 axis, a well-known pro-inflammatory signaling pathway^58,59^, mediates T cell and macrophage recruitment/migration in different tissues and diseases^60,61^ and promotes pro-inflammatory/pro-fibrotic M1 macrophage polarization^62^. To understand the functional relevance of our AS endometrial atlas, we incorporated scRNA-seq analysis into a phase I/II clinical trial of CD133+ BMDSCs in AS patients for the first time (**Supplementary Fig. 8A**). A comparison of endometrial cell transcriptomes from pre-treatment and post-treatment AS patients revealed that cell therapy reversed the appearance of AS-associated cell types, abnormal cell ratios, and aberrant CCC. Cell therapy in AS patients also increased the size of the main epithelial compartment while reducing the macrophage compartment. Differential gene expression analysis revealed an upregulation of genes related to angiogenesis (IGFBP2)^29^, epithelial development and cell proliferation (IGF1)^35^, and ECM remodeling (PDGFRA)^36^. Cell therapy also downregulated the expression of immune response- and chronic inflammation-related genes (MIF, IL6ST, DEFB1, and S100A10) and restored stromal and epithelial compartment communication through pathways related to epithelial differentiation and fibrosis (WNT)^51^ and cell proliferation and differentiation (NRG)^44^.

We also employed endometrial organoids as an in vitro model to recreate the endometrial epithelium of AS patients pre- and post-treatment with CD133+ BMDSCs, and compared them with healthy controls. As self-forming three-dimensional epithelial “mini-organs,” organoids reproduce the functionality and biology of epithelial compartments^63^. We observed a higher percentage of organoids generated from cells isolated from post-treatment AS patients during the first two passages (before equilibrating by passage 3), perhaps due to the restoration of epithelial cell fitness by CD133+ BMDSC treatment. Single-cell transcriptomic analysis of organoids identified primary in vivo epithelial populations: glandular, luminal, ciliated, and SOX9+ epithelium. We noted increases in SOX9+ and ciliated cell populations in post-treatment AS organoids, leading to a state resembling that of organoids from healthy endometrium. After cell therapy, these cell populations overexpressed the ECM glycoprotein (LAMC1), suggesting to play an important role in cell adhesion, differentiation, and proliferation^64^, and the inflammation-associated lncRNA NEAT1^31^.

In summary, we identified the differential endometrial cell niche of AS patients at single-cell resolution compared to normal endometrium during the secretory phase. Further, we report the partial reversion of abnormal cellular and transcriptomic composition following treatment with orphan-drug designated cell therapy using autologous CD133+ BMDSCs. Our in vivo and in vitro organoid models demonstrate the functional relevance of the AS endometrial cell atlas. Insights from this study may foster a better understanding of AS-associated pathological mechanisms and contribute to new therapeutic approaches targeting dysregulated cell-specific pathways.

## METHODS

### Subject details

The study was conducted in accordance with the International Conference on Harmonization Good Clinical Practice guidelines and the Declaration of Helsinki. All procedures involving human endometrium were approved by the Spanish Agency of Medicines and Medical Devices (Agencia Española de Medicamentos y Productos Sanitarios [AEMPS]) (April 20^th^, 2020) and the Clinical Research Ethics Committee at the Hospital Universitari Vall D’Hebron Barcelona, Spain (April 17^th^, 2020). All participants provided written informed consent. This study was registered at The European Union Clinical Trials Register (EU-CTR) and The Spanish Clinical Studies Registry (Registro Español de Estudios Clínicos [REec]) (EudraCT Number: 2016-003975-23; Registration date: April 21^st^, 2021; online: https://www.clinicaltrialsregister.eu/ctr-search/trial/2016-003975-23/ES; https://reec.aemps.es/reec/estudio/2016-003975-23). The Investigational Medical Product (IMP) was designed as Orphan Drug (OD) by the European Medicines Agency (EMA) on April 20^th^, 2017 (EMA/OD/313/16)^2^ and by the Food and Drug Administration (FDA) on February 1^st^, 2019 (designation request number DRU-2017-6131). For the design of this clinical trial, Scientific Advice and Protocol Assistance were conducted in 2017 with the AEMPS (February 10^th^) and the European Medicines Agency (EMA) (September 1^st^), respectively.

Nine patients of 34-42 years of age diagnosed with moderate and severe AS were enrolled according to the American Fertility Society (AFS) classification of IUAs^19^. The diagnosis of AS was confirmed by the same surgeon (X.S.), who performed all hysteroscopies and endometrial biopsies in the secretory phase immediately before and one month after BMDSC administration. All patients were treated with hormonal replacement therapy to synchronize cycles and treatments, and hysteroscopies were always performed during the WOI period. The endometrial microbiome was also assessed in patients with AS, which ruled out the existence of active endometritis that may affect typical endometrial tissue composition.

Requirements for participation in the study included a patient age of 18–44 years old, a BMI of 18-30, normal liver, heart, and kidney function, the absence of pregnancy, HIV, Hepatitis B or C, syphilis, or any psychiatric pathology, and a willingness to complete the study. BMDSC mobilization was performed using granulocyte-CSF (G-CSF) injection at 10 mcgr/kg. CD133+ cells were collected through peripheral blood apheresis and subsequently isolated. Patients were excluded if they did not reach the minimal requirements for mobilized cells (at least 30 million cells with a purity above 70% and viability >50%), presented an unstable medical condition, or refused a central venous catheter when required. Finally, isolated CD133+ cells were delivered into the spiral arterioles of the patient using a minimal invasive radiology intervention through the left brachial artery, reaching each uterine artery using a microcatheter (**Supplementary Fig. 5A**).

### Processing and dissociation of endometrial biopsies

A two-stage dissociation protocol separated endometrial biopsies into stromal fibroblasts and epithelial-enriched single-cell suspensions. Before dissociation, tissue was rinsed with PBS in a petri dish to remove blood and mucus, and excess PBS was removed after rinsing. The tissue was minced into small pieces and dissociated with 3 ml collagenase mix (Collagenase V, Sigma), RPMI+10% fetal bovine serum, and DNaseI (Sigma) at 37ºC under continuous shaking at 175 rpm for 25 min in a 15 mL Falcon tube in a horizontal position. This primary enzymatic step mostly dissociates stromal fibroblasts into single cells while leaving epithelial glands and lumen mostly undigested. The contents were transferred to a 50 mL tube in RPMI media and filtered with a 100 μm cell strainer. The tissue remaining on the filter was used for the epithelial enrichment cell isolation by resuspending and incubating in 10 ml Trypsin mix (Trypsin -EDTA (0.25%) phenol and DNaseI) for 10 min.

The resulting two contents were transferred to 50 mL tubes with 20 mL RPMI, filtered with a 100 μm cell strainer, centrifuged, and then resuspended in 1 mL RPMI. Dead cells were removed using the MACS dead cell removal kit (Miltenyi Biotec). Live-cell suspensions were loaded into a Chromium Next GEM Chip G (10X Genomics) to obtain cDNA from individual cells.

Mean cells obtained in stromal and epithelial enriched suspensions were 2.38 × 10^6^ (5 × 10^5^-6.4 × 10^6^) and 1.25 × 10^6^ (2 × 10^5^-5.3 × 10^6^), respectively. Viability of stromal cells was 67.44% (31-87%) and 39.5% (20-80%) for epithelial cells.

### 10x Genomics Chromium library preparation and sequencing

Total RNA was extracted using the RNeasy Micro Kit with on-column DNase treatment (Qiagen) following the manufacturer’s instructions. RNA was resuspended in 14 μl of RNase-free water, and purity and concentration determined using a NanoDrop 1000 spectrophotometer (Thermo Fisher). For cDNA synthesis, 500-1000 ng of total RNA was reverse transcribed following the manufacturer’s instructions (Thermo Fisher).

10x-Genomics v3 libraries were prepared as per the manufacturer’s instructions. Libraries were sequenced, aiming at a minimum coverage of 50,000 raw reads per cell, on an Illumina NovaSeq 6000 S2v1.5 300 cycles at a mean 40x coverage (paired-end; read 1: 26 cycles; i7 index: 8 cycles, i5 index: 0 cycles; read 2: 98 cycles)

### Alignment and quantification of the gene to cell count matrices from 10X scRNA-seq

Processing of scRNA-seq data was conducted with the CellRanger software suite (version 3.1.0). Raw reads were demultiplexed using the *mkfastq* wrapper command of *bcl2fastq* (Illumina). The libraries were mapped using GRCh38-3.0.0 as a reference provided by 10x Genomics. The count gene expression matrices per cell were computed per sample using the counts pipeline (with *--expect-cells* parameter set to default), which performs the steps of read alignment (with STAR mapping tool), UMI counting, calling of cell barcodes, and filtering of empty droplets based on a simple Good-Turing smoothing model of background gene expression profiles allows the discrimination of low RNA content cells from ambient RNA.

### Quality control filtering of cells and doublet detection

Low-quality cells in samples were filtered out using the distributions of the detected number of genes, detected number of counts, and mitochondrial percentage of read counts. Cells with 1 median absolute deviation (MAD) in at least two conditions were filtered out from the sample. These quality control and downstream analyses were performed in R version 4.1.

Doublet detection was performed with two methods: *DoubletFinder* (2.0.3 R package) and *scds* (1.6.0) R package. The expected doublet formation rate was fixed for each sample according to the cell count recovery given per sample by CellRanger. The hybrid approach from the *scds* package was used to avoid removing false-positive doublets. Cells marked as doublets by both algorithms were removed from the samples.

### Integration of scRNA-seq data across different conditions and cell clustering

Individual samples were aggregated, processed, and integrated with functions from the Seurat package (4.0.1). First, sample-to-sample aggregation for each condition dataset was performed with the *merge* function. Three aggregated datasets were constructed: (i) AS samples, (ii) patient-matched post-treatment AS samples, (iii) and control samples collected in our previous single-cell study of the natural menstrual cycle (GSE111976)^15^. The resultant count matrices were further processed to exclude cells with fewer than 750 detected genes and under a maximum mitochondrial content of 25%.

Each aggregated dataset was processed with the *SCTransform* function to integrate with the dataset of contrast to approach two integration analyses: (i) AS samples collapsed with control samples, and (ii) AS samples collapsed with matched cell therapy treated samples (further described in the methods sections). The *SCTransform* pipeline integrates different study conditions by applying a regularized negative binomial regression to allow the detection of condition-to-condition shared cell identities and the performance of differential expression analysis.

Mitochondrial ratios and cell cycle phase were regressed out from integrated datasets. *SelectIntegrationFeatures* was used to select the variable genes to integrate. *PrepSCTIntegration* and *FindIntegrationAnchors* were applied to identify the anchoring vectors across datasets and integration steps performed with *IntegrateData* (normalization method set to SCT). In the dimensional reduction step, the first 30 dimensions of PCA and UMAP were used to visualize cell clusters on the nearest-neighbor graphs.

Recoverable cells were manually labeled with the cell populations detected in AS when integrated into the post-treatment dataset to detect cell types unique to the disease condition. These labels were downstream transferred to the control dataset by the anchor integration protocol in the following analysis steps.

### AS cartography: Integration by the dataset of origin

Control samples were retrieved from the 10X Single-cell 3‘v3.1 dataset (available at GSE111976). Cell populations detected in AS were aggregated with retrieved control cells. The Seurat Integration protocol was applied to integrate the two datasets. Raw Seurat objects of control and AS samples were split into two objects. The *SCTransform* function processed each object to processing counts. The dataset was further integrated using the 3000 most variable genes. *IntegrateData* was applied using the dataset of origin as an integration factor. Validation of the integration step was assessed by the evaluation of the conservation of canonical markers across AS and control conditions (**Supplementary Fig. 1B** and **Supplementary Table 2**)

### Single-cell mapping during cell therapy: Integration according to treatment

Pre- and post-treatment AS samples were similarly integrated using the same SCT pipeline. Non-processed Seurat objects from pre-treatment and post-treatment datasets were split into separate objects based on the treatment stage.

In this case, the 4000 most variable genes present in combined datasets using the *SelectIntegrationFeatures* function were used. The *clustree* (0.4.4) package was used to evaluate the optimal resolution number in the *FindCluster* function; multiple clusters with a range of resolution from 0.1 to 2 were generated, with increased steps of 0.1. A manual review of the generated UMAPs and *clustree* output determined an optimal resolution of 0.8 to label the different clusters. Validation of the integration step was assessed by evaluating the conservation of canonical markers across pre- and post-conditions (**Supplementary Fig. 9C** and **Supplementary Table 4**).

### Annotation of cell types in scRNA-seq datasets

Cell type identification was conducted to examine each cluster’s specific differentially expressed genes compared to other clusters. The Wilcoxon Rank Sum test was used, and the p-values were adjusted using the false discovery rate (FDR) method. Cluster cell identity was evaluated by comparison of cluster marker genes with available biological knowledge. Differentially expressed marker genes for each cell type are available in **Supplementary Table 1**.

### Analysis of significant changes in cell population sizes

Cell ratio analysis across different experimental groups was performed using a pseudobulk strategy. A count matrix of the different cell types detected per sample was created, and the *edgeR* (3.34.0) package was used to compute a differential abundance (DA) test across cell types. The FDR p-value adjusting method was applied. Cells detected as cycling cells were removed prior to DA analysis. All cell type ratios of AS vs. Control and pre-treatment vs. post-treatment comparisons are available in **Supplementary Tables 8** and **9**, respectively.

### Differential expression and gene functional enrichment analysis

The *FindMarkers* function from the *Seurat* package was used to perform differential expression analysis across the treatment stages (**Supplementary Tables 3** and **5**) and across the different cell types reported in the dataset (**Supplementary Tables 1, 2**, and **4**). The Wilcoxon Rank Sum test was used, and the p-values were corrected using the FDR method. Genes with FDR values under 0.05 were considered significant. GSEA was performed using *escape* (1.6.0) R package^65^ to study the menstrual cycle function in the secretory phase between treatment groups. The *enrichIt* function was used to compute the score for each cell present in the dataset, with the *UCell* method to compute the scores. Finally, the score differences between treatment stages in the endothelium, epithelium, AS epithelium, perivascular, and stromal cell fractions were evaluated. The *getSignificance* function was used to perform the statistical testing using a Wilcoxon test; p-values were corrected using the FDR method (**Supplementary Table 6**). The following gene set related to menstrual cycle function in the secretory phase was used^15^: SCGB1D2, MT1F, MT1X, MT1E, MT1G, CXCL14, WNT5A, SFRP1, ZFYVE21, CILP, SLF2, MATN2, and S100A4.

### CCC network analysis

Gene expression and metadata matrices were used as input for the *CellChat* (1.1.3) package to infer the potential autocrine and paracrine interactions between cell populations and apply differential CCC network analysis for AS vs. Control, and pre-treatment vs. post-treatment experimental groups. Only common cell types can be included in group comparisons. The influence of each cell population was corrected using the number of cells detected in each group by activating the argument *population*.*size* in the function *computeCommunProb*. Cells labeled as *cycling* were removed from downstream CCC analysis. The CCCs were also filtered to eliminate those with less than ten cells. Finally, the *ranknet* function was applied for network differential testing; a p-value threshold of 0.01 was applied to consider significant changes between groups.

### Organoid culture from endometrial biopsies

Organoids were generated from AS patients’ endometrium pre- and post-treatment and from healthy control endometrium. After enzymatic dissociation of the endometrial biopsies (described above), epithelial and stromal cells were obtained (in the same tube), and total cell numbers were counted. After centrifugation at 300 g, the pellet was resuspended in 70% Matrigel (Corning, 356231) and 30% DMEM/F12 (Thermo Fisher Scientific, 11330032) and supplemented with the ROCK inhibitor Y-27632 (1/500). The suspension was cultured in 20-µl droplets containing 25,000 cells, deposited in prewarmed 48-well plates, and allowed to form a gel at 37ºC and 5% CO2 before adding culture medium. Organoid culture medium was prepared as previously described^16,66^ (**Supplementary Table 7**). Matrigel droplets were cultured for 14-16 days in the first passage (P0) with medium changes every two days. Organoids were then grown for seven days before the following passage, with media changes every two days.

Organoids were recovered for passaging by liquifying the Matrigel droplets with ice-cold DMEM/F12 (without any growth factors, serum, or enzymes), followed by repeated pipetting to ensure maximum recovery. Organoids were then dissociated by incubation with TrypLE supplemented with Y-27632 (1/1000) and mechanically triturated. The resulting cell suspension was centrifuged at 300 g, and the pellets were resuspended in 70% Matrigel and 30% DMEM/F12 supplemented with Y-27632 and placed as droplets in 48-well plates. Two passages (till P2) were performed to remove the contaminating stromal cells from the epithelial organoids before scRNA-seq analysis. After seven days of P2 culture, organoids were dissociated for single-cell transcriptomic analysis following the same protocol described for passaging but with an additional cell counting step for the single-cell transcriptomics protocol (10x Genomics, described above).

### Organoid counting

Multiple microscopy images (Thermo Fisher, EVOS M5000) were taken (for a reliable representation of total organoids) to count organoids present in each Matrigel droplet. In particular, images were taken by dividing each drop into two depth sections and each section into four regions. Two images were counted diagonally from each section, and the mean of these images was calculated. This mean was multiplied by the number of divisions of each section (4) and the number of sections (2) to infer the total organoid number per drop. For each condition, two experimental replicates were measured.

### Identifying organoid-resident cells using machine learning

The transcriptomic identity of each cell relative to in-vivo cell subtypes was profiled to analyze organoid-derived cells following a similar approach as that described by La Manno et al.^67^. Detected epithelial cells from the GSE111976 dataset were used, and the different subpopulations were studied. Previously described gene cell markers were used to label cells^15,20^.

Before training the model, a feature selection of genes was performed. First, the top 4000 most variable genes were detected using the *FindMarker* function from Seurat. Second, the specific gene markers of each cell type were computed using only the previously selected gene set. Finally, the top 1000 genes ranked by three metrics of cell type specificity were selected (fold-increase, fold-increase*fraction-positive, and fold-increase*fraction-positive^0.5). A machine learning logistic regression model (from *caret* (6.0-92) and *glmnet* (4.1-4) R packages^68^ was then used to predict the identity of organoid cells. The training dataset was split randomly, with 70% of cells used for training and 30% for evaluation in the testing phase. Fine-tuning of the logistic regression hyperparameters was carried out as follows: cross-validation of ten iterations with the lambda value changed from 0.001 to 0.01 in steps of 0.01. Alpha was held constant at a value of 1. The optimal lambda value was 0.001, and a 0.9446 accuracy (0.9402, 0.9488 95% CI) in the test phase was achieved. Finally, the probabilities of the inferred cell types of organoids were plotted in a polygonal projection.

Finally, the differences between cell type probabilities across experimental groups were evaluated by applying the Kruskal–Wallis test following Dunn’s multiple comparison test. P-values were corrected using the Bonferroni method. Differential expression in organoid cells between AS-pre and AS-post was assessed by applying the Wilcoxon Rank Sum test (FDR<0.05) as previously described. Results are supplied in **Supplementary Table 10**.

## Data Availability

All data produced in the present study are available upon reasonable request to the authors.

## DATA AVAILABILITY

The single-cell RNAseq data generated for this manuscript has been uploaded to GEO, under accession number GSE215968 (https://www.ncbi.nlm.nih.gov/geo/query/acc.cgi?acc=GSE215968). This dataset includes endometrial samples of Ashermans’ Syndrome pre-treated and post-treated patients, and their matched authologous BMDSCs CD133+ sorted samples. Control samples of 10 healthy endometrial biopsies were downloaded from GEO: GSE111976 (PMID: 32929266).

Other data that support the findings of this study are available from Asherman Therapy SL, but restrictions apply to the accession of these data, which were used under license for the current clinical study, and so are not publicly available. Data are however available from the authors upon reasonable request and with permission of Vall Hebron Ethical Committe.

## CODE AVAILABILITY

Not applicable.

## ACKNOWLEDGEMENTS

We thank Dr. Nicolas Plachta, University of Pennsylvania for valuable discussions and advice. This study was jointly supported by Human Uterus Cell Atlas Project from the European Union’s Horizon 2020 research and innovation programme under grant agreement No. 874867, PROMETEO/2018/161 from the Valencia Government, IDI-20201142 CDTI from the Spanish Government and Carlos Simon Foundation, Spain.

B.R. was supported by the H2020 funded project Human Uterus Cell Atlas (HUTER) (2020/2021) (Grant Agreement 874867). R.P. was supported by an Industrial Doctorate grant, (DIN2020-011069) from the Spanish Ministry of Science and Innovation (MICINN). N.K. was supported by PROMETEO/2018/161. J.G.F. was support by PFIS grant [FI19/00159]. I.M. was supported by a FIS project grant [PI21/00235]. F.V. was supported by a FIS project grant [PI21/00528].

## AUTHOR CONTRIBUTIONS

X.S., B.R., F.V., I.M., and C.S. conceived and designed the study. J.G.-F., N.K. and E.F. performed single-cell experiments. X.S. performed the clinical trial EudraCT Number: 2016-003975-23 and collected endometrial biopsies. J.G.-F and H.V performed organoid experiments. B.R. and R.P analyzed single-cell RNA-seq data. X.S., B.R., F.V., I.M., and C.S. interpreted the results and wrote the manuscript.

## COMPETING INTERESTS

C.S. & X.S are founders and shareholders of Asherman Therapy S.L. The rest of authors declare no competing interests.

SUPPLEMENTARY MATERIAL

**Supplementary Figure 1. A)** Dotplot showing top 5 gene markers that allow the identification of cell types in the AS integrated atlas. Full table of DE genes is included in Supplementary Table 1. **B)** Dotplot showing top 5 conserved genes between AS and control endometrial cell types. Full list of genes is included in Supplementary Table 2. **C)** Cell ratios comparing the endometrium of AS patients and healthy subjects (Control). **D)** Marker gene expression in unique AS-related cell types (AS Epithelium and SMC cells). **E)** Integrated UMAP with highlighted unique AS types. Abbreviations: AS: Asherman; DC: dendritic cells; DN: double negative; MAIT: mucosal-associated invariant T; NK: natural killer; PV: perivascular; SMC: smooth muscle cells; UMAP: Uniform manifold approximation and projection.

**Supplementary Figure 2**. Differential expression between AS and control samples separated by cell type.

**Supplementary Figure 3. A)** CCC network showing significant ligand-receptor interactions in AS vs Control samples. **B)** Heatmap of the overall signaling patterns between cell types in AS and Control groups. Abbreviations: AS: Asherman; CCC: Cell-to-cell communication; DC: dendritic cells; DN: double negative; MAIT: mucosal-associated invariant T; NK: natural killer; PV: perivascular; SMC: smooth muscle cells.

**Supplementary Figure 4**. Dotplot of CCC results of COLLAGEN signaling pathway. (AS in red labels, Control in green labels).

**Supplementary Figure 5**. Dotplot of CCC results of fibronectin (FN1) signaling pathway. (AS in red labels, Control in green labels).

**Supplementary Figure 6**. Dotplot of CCC results of LAMININ signaling pathway. (AS in red labels, Control in green labels).

**Supplementary Figure 7**. Dotplot of CCC results of CCL signaling pathway. (AS in red labels, Control in green labels).

**Supplementary Figure 8. A)** Overview of study design and instillation of CD133+ cell therapy with microcatheter in the right and left spiral arteries of the uterus. **B)** Violin plot of the expression of PROM1 across patient samples in CD133+ cluster. **C)** UMAP of 77,700 autologous FACS-sorted CD133+ cells from ten AS patients. **D)** UMAP plot and expression of PROM1 in different clusters detected. **E)** Dotplot of gene markers that distinguish cell clusters and PROM1 expression.

**Supplementary Figure 9. A)** Yield of endometrial cells per patient before and after CD133+ BMMDCs injection. **B)** Boxplot of cell type ratios comparing AS patients at pre-treatment (AS-Pre) and post-treatment (AS-Post) stages. **C)** Dotplot showing top 5 gene markers conserved between pre-treatment and post-treatment conditions. **D)** Overlap of cell clusters by treatment condition and **(E)** by patient.

**Supplementary Figure 10**. Differential expression between AS-Pre and AS-Post in different cell types. Abbreviations: DC: dendritic cells; DN: double negative; NK: natural killer; PV: perivascular.

**Supplementary Figure 11**. Dotplot of CCC results of FGF signaling pathway (AS-Pre in red labels, AS-Post in blue labels).

**Supplementary Figure 12. A)** Cell-to-cell communication chord plots in AS-Pre and AS-Post stages of LIGHT signaling pathway. **B)** Cell-to-cell communication chord plots in AS-Pre and AS-Post stages of ICAM signaling pathway. **C)** Cell-to-cell communication chord plots (upper panel) and pathway interactor molecules (lower panel) of IL4 pathway in AS-Pre stage. **D)** Cell-to-cell communication chord plots (upper panel) and pathway interactor molecules (lower panel) of RESISTIN signaling pathways in AS-Pre stage.

**Supplementary Figure 13. A)** Scheme of organoid generation and analysis (Control (n=3), AS (n=3, paired pre-treatment and post-treatment). **B)** Number of organoids counted in different passages for AS patients’ pre-treatment and post-treatment. **C)** Representative images of organoid culture generated from patient’s 2 samples pre-treatment and post-treatment at different passages (P0= Passage 0, P1= Passage 1, P2= Passage 2) and controls. **D)** Single-cell UMAP distribution of control freshly isolated cells used as training set for the logistic regression model to predict cell types in organoid cells. **E)** Histograms displaying the prediction score of organoid cells over in vivo reference cells.

**Supplementary Table 1**. Cell type marker genes.

**Supplementary Table 2**. Conserved marker genes between AS and control conditions.

**Supplementary Table 3**. Differentially expressed genes between AS and control conditions.

**Supplementary Table 4**. Conserved marker genes between pre-treatment and post-treatment conditions.

**Supplementary Table 5**. Differentially expressed genes between pre-treatment and post-treatment conditions.

**Supplementary Table 6**. Enrichment results on secretory phase genes.

**Supplementary Table 7**. Organoid’s culture medium.

**Supplementary Table 8**. Cell ratios between AS and control conditions.

**Supplementary Table 9**. Cell ratios between pre-treatment and post-treatment conditions.

**Supplementary Table 10**. Differentially expressed genes in organoid cells.

