## Supplementary figure 7 for "Decoding the endometrial niche of Asherman’s Syndrome at single-cell resolution"

**Supplementary Table 7. Organoid’s culture medium.**

| **Reagent** | **Concentration** | **Commercial brand** |
| --- | --- | --- |
| DMEM F12 (+Glutamine +HEPES) | Fill to the volume needed | Thermo Fisher Scientific |
| Noggin | 100 ng/ml | R&D Systems |
| RSPO-1 (R-spondin 1) | 200 ng/ml | R&D Systems |
| Insulin-Transferrin-Selenium (ITS) | 1% | Life Technologies |
| Penicillin/streptomycin | 1% | Life Technologies |
| N2 supplement | 1% | Life Technologies |
| B27 supplement | 2% | Life Technologies |
| N-acetyl L-cysteine | 1.25 mM | Merck |
| Nicotinamide | 1 mM | Merck |
| A83-01 | 0.5 µM | Merck |
| p38 inhibitor (SB202190) | 10 µM | Merck |
| EGF (epidermal growth factor) | 50 ng/ml | R&D Systems |
| b-FGF (basic fibroblast growth factor) |  |  |
| FGF10 (fibroblast growth factor 10) | 50 ng/ml | Peprotech |
| Rock Inhibitor | 9 µM | Merck |
